# Data-driven analysis on the simulations of the spread of COVID-19 under different interventions of China

**DOI:** 10.1101/2020.05.15.20103051

**Authors:** Ting Tian, Jingwen Zhang, Shiyun Lin, Yukang Jiang, Jianbin Tan, Zhongfei Li, Xueqin Wang

**Affiliations:** School of Mathematics, Sun Yat-sen University, Guangzhou, 510275, China; School of Management, University of Science and Technology of China, Hefei, 230026, China; School of Management, Sun Yat-sen University, Guangzhou, 510275, China

**Keywords:** Pre-symptomatic transmission, Bayesian inference, Latent Markov Model, Time-varying reproduction number, Mode migration, Policy intervention

## Abstract

Since February 2020, COVID-19 has spread rapidly to more than 200 countries in the world. During the pandemic, local governments in China have implemented different interventions to efficiently control the spread of the epidemic. Characterizing transmission of COVID-19 under some typical interventions is essential to help countries develop appropriate interventions. Based on the pre-symptomatic transmission patterns of COVID-19, we established a novel compartmental model: Baysian SIHR model with latent Markov structure, which treated the numbers of infected and infectious individuals without isolation to be the latent variables and allowed the effective reproduction number to change over time, thus the effects of policies could be reasonably estimated. By using the epidemic data of Wuhan, Wenzhou and Shenzhen, we migrated the corresponding estimated policy modes to South Korea, Italy, and the United States and simulated the potential outcomes for these countries when they adopted similar policy strategies of three cities in China. We found that the mild interventions implemented in Shenzhen were effective to control the epidemic in the early stage, while more stringent policies which were issued in Wuhan and Wenzhou were necessary if the epidemic was more severe and needed to be controlled in a short time.

## 1. Introduction

Since December 2019, the report of COVID-19 detected was sent to the World Health Organization (WHO) by Chinese government [25]. China issued a series of intervention policies to reduce the transmission of COVID-19, such as suspending public transportations in areas of severe outbreak (e.g. Wuhan), cancelling public gatherings and delaying reopening of enterprises. These endeavours have led to the significant suppression of the epidemic of COVID-19. There was no domestically newly reported confirmed cases for the first time in China on March 18, 2020 [27]. The local and national responses of China have made remarkable contributions to the prevention and control of COVID-19.

In this article, we selected Wuhan, Wenzhou and Shenzhen cities as the typical representatives of the policy interventions of China. Wuhan was the most severe epidemic city of COVID-19 outbreak in China. It suspended public transportation, cancelled all outbound trains and flights[23] and started to construct a specialist emergency hospital (i.e. Huoshenshan Hospital) for patients on January 23, 2020. Wenzhou was the city with the largest number of cumulative confirmed cases in Zhejiang province and implemented comparatively stringent policy interventions for the control of COVID-19. It announced that every household might send one person every two days outside for necessity purchases[24]. Shenzhen was the city of the confirmed cases initially reported in Guangdong province, the intervention policy was relatively moderate due to its special economic status in China. It announced that all the individuals returning from Hubei needed to be isolated at home or other suitable facilities for 14 days on February 2, 2020[21], and the residential units with confirmed cases were forced to implement “hard quarantine” for 14 days. All details about the interventions implemented in these cities were given in Figure 1.

**Figure 1.**
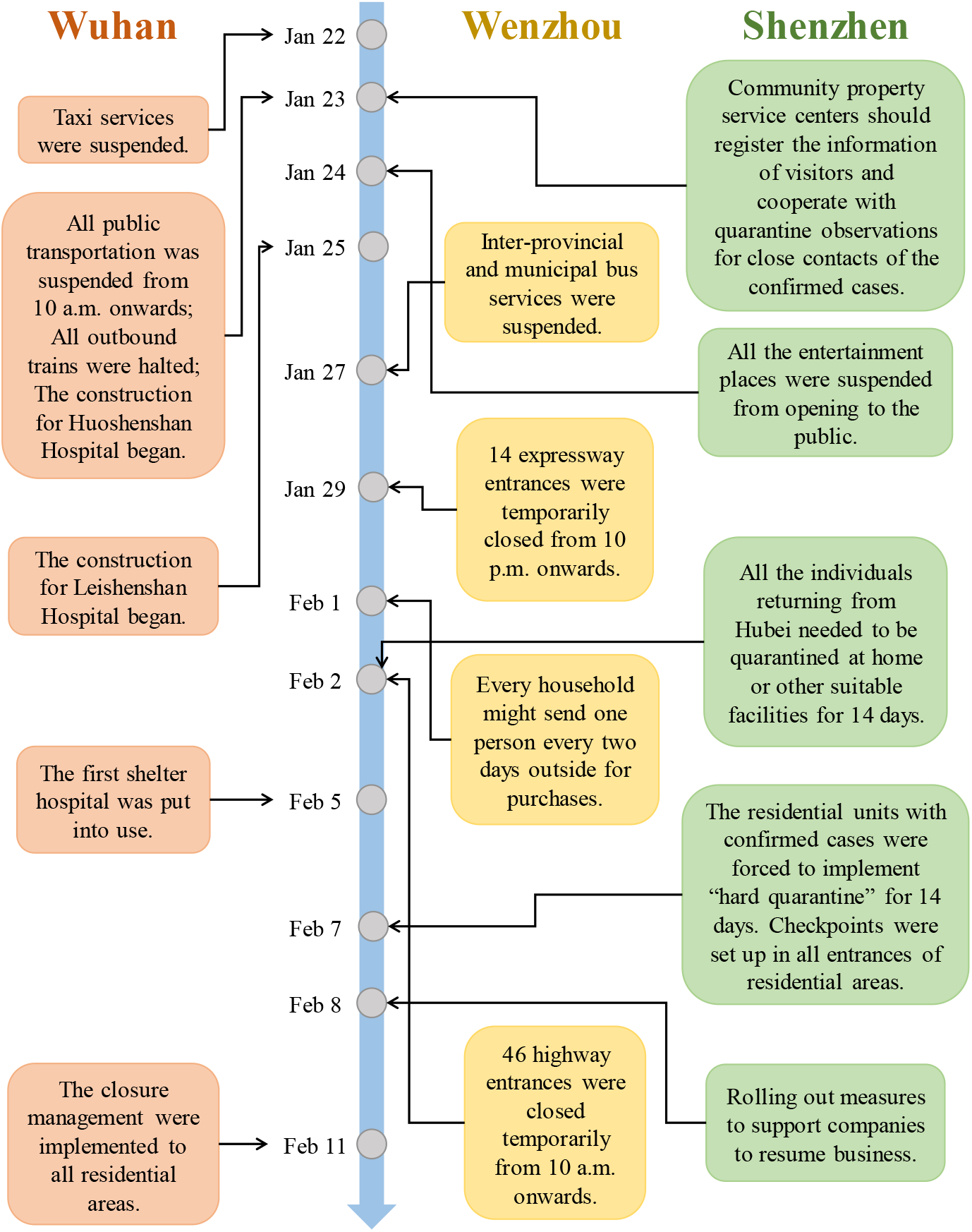
The timeline of interventions implemented in Wuhan, Wenzhou, and Shenzhen.

Nevertheless, these policy interventions were implemented at the expense of the economic loss. Taking individual consumption industry as an example, the amount of total retail sales of consumer goods decreased by about 20.5% with a sharp drop of catering services by 43.1% from January to February 2020 compared to the same duration of 2019 according to the National Bureau of Statistics in China [17]. Consequently, policymakers needed to anticipate the likely outcomes of interventions in terms of their own epidemic situations.

The epidemic has rapidly spread to more than 200 countries around the world. WHO declared that COVID-19 became a pandemic on March 11, 2020 [26]. As an epitome of the pandemic, the numbers of cumulative confirmed cases and deaths on May 31, 2020 were 11468 and 270 in South Korea, 232664 and 33340 in Italy, and 1819788 and 105634 in the United States, respectively[1]. Some details about the spread of COVID-19 in South Korea, Italy and the United States are given in COVID-19 prevalence in Supplemental material. In this situation, the control measures of China, which have significantly suppressed the spread of COVID-19, are worth learning for other countries confronting with coronavirus. Therefore, it would be instructive to simulate the potential outcomes for other countries when they adopted similar policy strategies of China.

In order to simulate the likely outcomes under different policy patterns of China for other countries reasonably, an epidemic model reflecting the nature of COVID-19 and the effects of policy interventions is needed. Based on the report of WHO[9], the transmission of COVID-19 could be caused by the individuals infected with the virus before significant symptoms developed. Because of the lacking assumption of incubation period, traditional compartmental model, such as SIR model[5, 7], were inappropriate to be used in the evaluation of the spread of COVID-19. Moreover, although the typical SEIR model [14, 15] took the effects of incubation period into account, it needed some modifications of the delay effects [12, 28] for considering pre-symptomatic transmission of COVID-19, which was subjective and indirect for reflecting the transmission patterns of COVID-19.

In this paper, we proposed a novel compartmental epidemic model based on P-SIHR Probabilistic Graphical Model[22]: Bayesian SIHR model, which considers presymptomatic transmission of COVID-19 by an appropriate compartmental assumption with the well-defined time-varying reproduction number. In Bayesian SIHR model, the numbers of infected and infectious individuals without isolation are the key ingredients for the evaluation of the spread, which are assumed to be latent variables correlated with numbers of compartments at different time by a latent Markov structure. We used the estimated time-varying reproduction number[11, 20] to reflect the effects of interventions implemented in Wuhan, Wenzhou, and Shenzhen, and then migrated the corresponding policy patterns by parameters transition to simulate the likely outcomes of these interventions in South Korea, Italy and the United States, so that the lessons can be learned from the key decisions made in the representative cities.

The rest of this paper is organized as follows. In Section 2, we introduce the construction of our proposed model, describe the parameter estimation procedure in details and further show the mode migration approaches in policy pattern assessment. In Section 3, we present the simulation study for the spread of COVID-19 in South Korea, Italy and the United States under different interventions of China by the proposed model. Finally, we end up with conclusions in Section 4.

### 2. Epidemic Modelling

#### 2.1. Data sources

The epidemic data were extracted for Wuhan, Wenzhou, and Shenzhen from the Chinese Center for Disease Control and Prevention (China CDC) [18], we used the reported numbers of cumulative confirmed cases, cumulative recovery cases and total deaths from January 15 to February 12 in Wuhan, from January 21 to February 9 in Wenzhou and from January 19 to February 6 in Shenzhen. Similarly, the reported numbers of cumulative confirmed cases, cumulative recovery cases and total deaths for South Korea, Italy and the United States were downloaded through April 23, 2020 from “nCov2019” package [1].

#### 2.2. Bayesian SIHR model

To understand the pre-symptomatic transmission patterns of COVID-19, we introduced Susceptible-Infected and infectious without isolation-Hospitalized in isolation-Removed (SIHR) model with four compartments: Susceptible (*S*), infected and infectious without isolation (*I*), hospitalized in isolation (*H*), removed (*R*) and assumed that:

1. The susceptible (*S*) individuals have no immunity to the disease;
2. The infected and infectious individuals without isolation (*I*) were assumed to be infectious, transmissible and would transform into the hospitalized in isolation eventually after symptoms onset;
3. The hospitalized (*H*) individuals were in isolation and treated carefully and would not transmit COVID-19 to the susceptible;
4. The removed (*R*) individuals would not be infected again.

Based on the above assumptions, the transfer relationships between compartments *S, I, H, R* were given in Figure 2:

**Figure 2.**
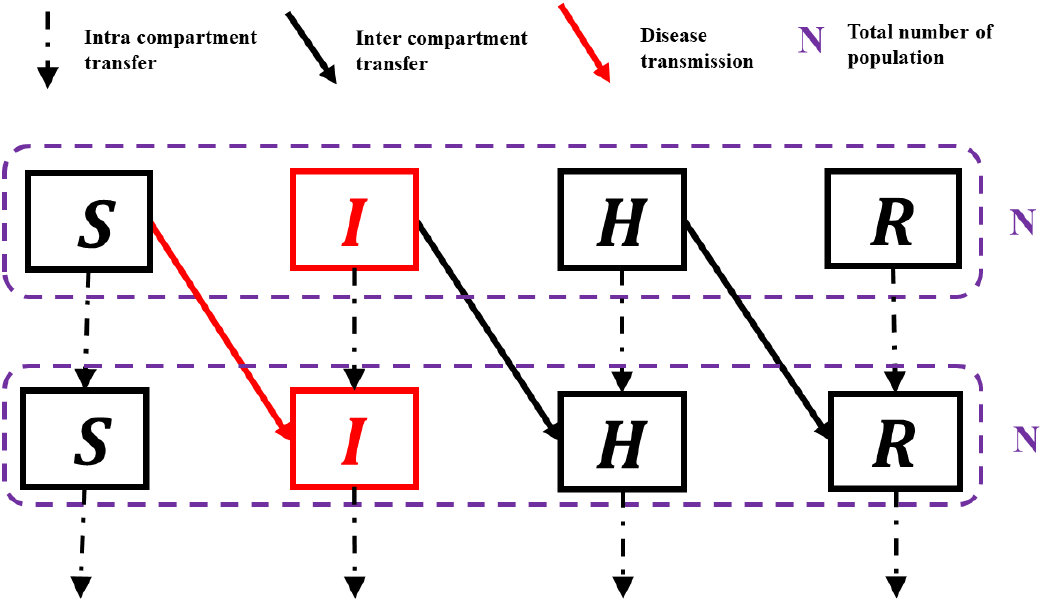
The diagram of transfer process of four compartments. Susceptible (*S*), infected and infectious without isolation (*I*), hospitalized in isolation (*H*), and removed (*R*).

Notice that an increase in the number of infected individuals only happens through the contact between *S* and *I*, similar to SIR model [5, 7], we can define the corresponding dynamic system of these four compartments as follows:

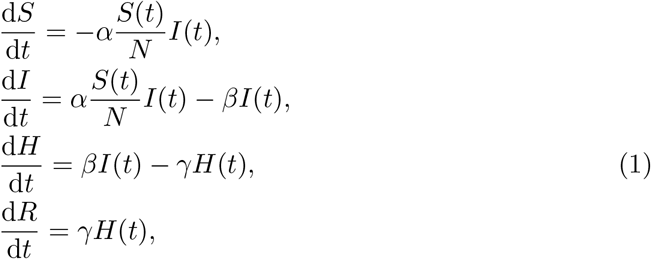

where (*S*(*t*), *I*(*t*), *H*(*t*), *R*(*t*)) are the numbers of corresponding compartments at time *t*. Conatct rate *α* denotes the average contact numbers per person during the outbreak of infectious disease, 1*/β*, 1*/γ* are the average lengths of retention time of a person for compartments *I* and *H*, respectively, where 1*/β* is the mean of the length of incubation period of COVID-19, and is assumed to be 5 days in terms of studies about COVID-19 [3, 10, 16].

The basic reproduction number *R*_0_, which is the expected number of secondary cases produced by a single (typical) infection in a completely susceptible population, is defined as the spectral radius of the next generation operator [11, 20] of SIHR model:

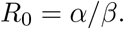

Taking policy interventions into consideration, it’s more reasonable to assume the time-varying transmission patterns in SIHR model, which can be achieved by the assumption of time-varying reproduction number *R*_*t*_ [8].

To obtain *R*_*t*_, we supposed that the parameter *α* was time variant and would decrease with the interventions. We used logistic function [22] to simulate the decreasing trend of *α*:

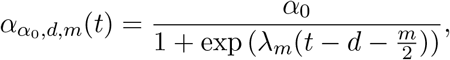

where *α*_0_ denotes the maximum contact numbers per person during the early outbreak of infectious disease, *d* is the time when the control measures start to be effective and *α*_0_ starts to decline, *m* represents the duration of a process where the epidemic is to nearly vanish, *λ*_*m*_ is chosen as 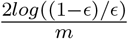 and *ϵ* is fixed to be 0.01. The smaller the values of *d* and *m*, the earlier effectiveness and the stronger intensity of interventions were implemented, respectively. The graph of time-varying contact rate *α* is given in Figure 3:

**Figure 3.**
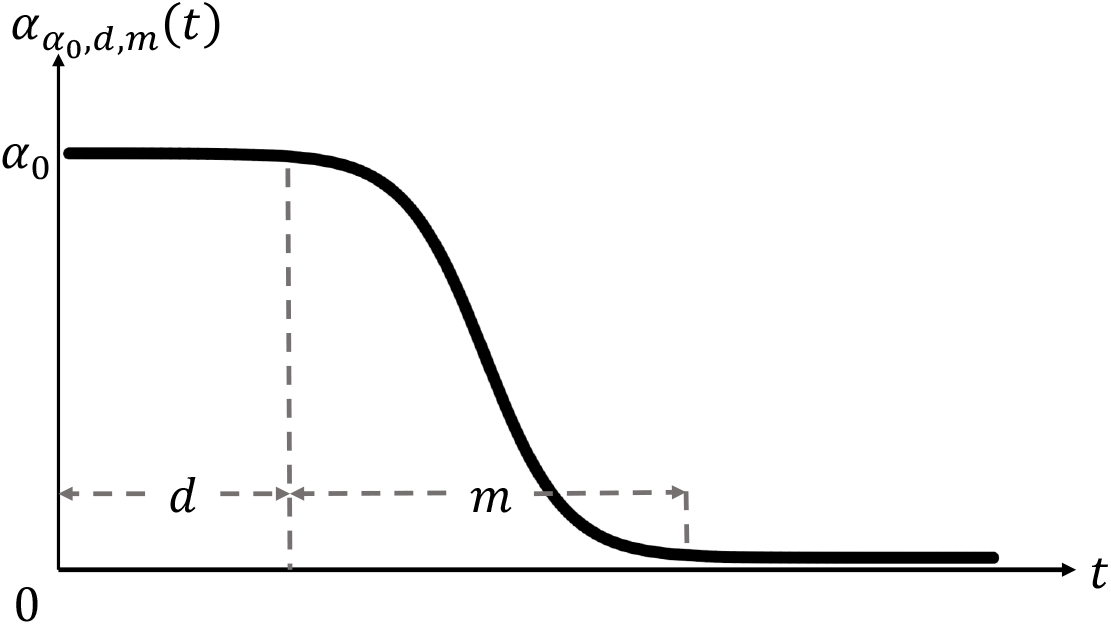
The time-variant contact rate *α*

By the setting above, the corresponding dynamic system of SIHR model with time-varying reproduction number along with parameters *α*_0_, *d, m, β, γ*, were given as follows:

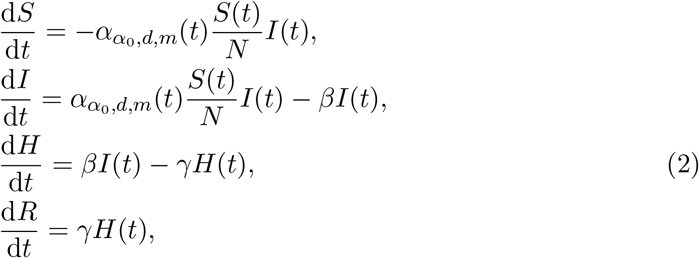

where the corresponding *R*_*t*_ was defined as:

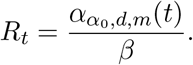

For simplicity, let 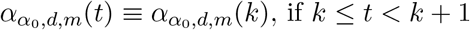.

While time-varying SIHR model defined above could be effective to consider the pre-symptomatic transmission patterns of COVID-19 and the effects of policy interventions, in order to fit the observed data appropriately, we introduced randomness to time-varying SIHR model by the latent Markov structure: let Ω := (*α*_0_, *d, m, γ*), which are the parameters needed to be estimated under the given *β*, and *S*_*t*_, *I*_*t*_, *H*_*t*_, *R*_*t*_ be the random numbers of compartments *S, I, H, R* at time *t. A*_*t*_ := (*S*_*t*_, *I*_*t*_, *H*_*t*_, *R*_*t*_) is assumed to be a multidimensional Markov process under constraints *N* = *S*_*t*_ + *I*_*t*_ + *H*_*t*_ + *R*_*t*_, i.e. for *g < t*,

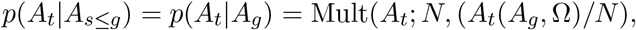

where *A*_*t*_(*A*_*g*_, Ω) is the vector of the numbers of compartments *S, I, H, R* at the time *t* determined by the deterministic dynamic system (2) given the initial value *A*_*g*_ and parameters Ω and *β*. Mult(· ; *n, p*) is the density of Multomial distribution with the total number *n* and the incident rate vector *p*. and.

Notice that the observable data are the numbers of cumulative confirmed cases (*c*_*t*_), cuMultative recovery cases (*r*_*t*_) and total deaths (*d*_*t*_) at each time *t*. According to the definition of compartments *R* and *H*, we have *R*_*t*_ = *r*_*t*_ + *d*_*t*_ and *H*_*t*_ = *c*_*t*_ *− R*_*t*_. Based upon the above assumptions, the observed data for stochastic time-varying SIHR model are *H*_1:*T*_, *R*_1:*T*_, *N*, where *T* is the observed length of time. We treated *I*_1:*T*_ as the latent variables, then the randomness of *S*_*t*_, *I*_*t*_, *H*_*t*_,and *R*_*t*_ construct a latent Markov structure for the dynamic process of epidemic.

#### 2.3. The estimation procedure

We employed Bayesian framework [6, 13] for the statistical inference of Ω and *I*_1:*T*_. Once the prior distribution of Ω: *π*(Ω) were given, we used the posterior distribution of Ω and *I*_1:*T*_ : *π*(Ω, *I*_1:*T*_ |*H*_1:*T*_, *R*_1:*T*_, *N, β*) for evaluating the dynamic process of epidemic. Notice that:

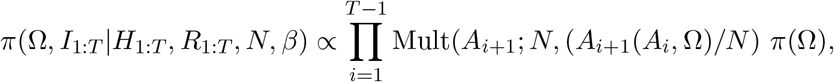

Considering the fact that: *S*_*t*_ ≫ *I*_*t*_, *H*_*t*_, *R*_*t*_. If the policy interventions are implemented to suppress the spread of infectious disease, we have:

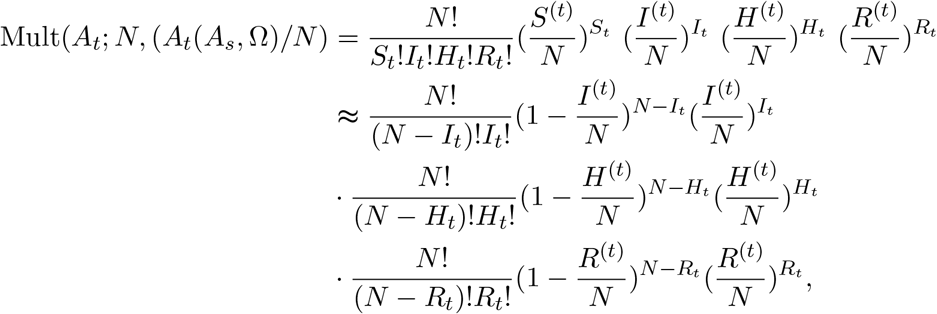

where (*S*^(*t*)^, *I*^(*t*)^, *H*^(*t*)^, *R*^(*t*)^) := *A*_*t*_(*A*_*s*_, Ω).

If *N → ∞*, we can use Poison approximation for the marginal distributions of *I*_*t*_, *H*_*t*_, *R*_*t*_, which implies that *I*_*t*_ *∼* Pois(*I*^(*t*)^), *H*_*t*_ *∼* Pois(*H*^(*t*)^), *R*_*t*_ *∼* Pois(*R*^(*t*)^) and *S*_*t*_ = *N − I*_*t*_ *− H*_*t*_ *− R*_*t*_ given *A*_*s*_, where Pois(*λ*) means the Poison distribution with mean *λ*. We used this approximation for the posterior distribution *π*(Ω, *I*_1:*T*_|*H*_1:*T*_, *R*_1:*T*_, *N, β*). Since *π*(Ω, *I*_1:*T*_007C*H*_1:*T*_, *R*_1:*T*_, *N, β*) has no closed form, combining with flat prior for Ω, we used Gibbs sampler embedded Metropolis-Hastings steps [2, 4] to sample *π*(Ω, *I*_1:*T*_ |*H*_1:*T*_, *R*_1:*T*_, *N, β*), the full conditional distributions of Ω and *I*_*t*_ are as follows:

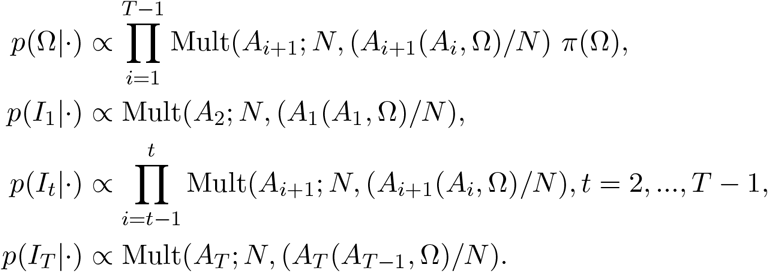

The technical problem within the iteration of Markov chain Monte Carlo (MCMC) is the sampling of latent variables *I*_1:*T*_, which is high dimensional if *T* is large. For sampling *I*_1_, note that at the early stage of epidemic, *S*_1_ *≈ N*, which implies that

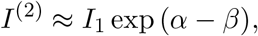

where (*I*^(2)^, *H*^(2)^, *R*^(2)^) := *A*_2_(*A*_1_, Ω), then the full conditional distribution of *I*_1_ approximately achieves:

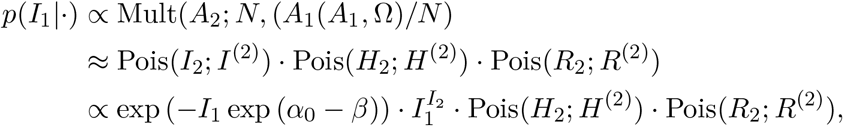

which means that we can use Gamma(*I*_2_ + 1, exp (*α*_0_ *− β*)) as the proposal transfer kernel of *I*_1_ for well approximation of *p*(*I*_1_|·). Given the sample of *I*_1_, there are two strategies for the sampling of *I*_2:*T*_ :

1. Group sampling: For *i* in 2 to *T, I*_*i*_ *∼* Pois(*I*^(*i*)^) given Ω, (*S*_*i−*1_, *H*_*i−*1_, *R*_*i−*1_) and the sample *I*_*i−*1_, and then accept the whole *I*_2:*T*_ with given probability in Metropolis-Hastings steps, where (*S*^(*i*)^, *I*^(*i*)^, *H*^(*i*)^, *R*^(*i*)^) := *A*_*i*_(*A*_*i−*1_, Ω).
2. Sequential sampling: For *i* in 2 to *T, I*_*i*_ *∼* Pois(*I*^(*i*)^) given Ω, (*S*_*i−*1_, *H*_*i−*1_, *R*_*i−*1_) and the accepted sample *I*_*i−*1_ in (*I −* 1)-th step, and then accept the *I*_*i*_ with given probability in Metropolis-Hastings steps, where (*S*^(*i*)^, *I*^(*i*)^, *H*^(*i*)^, *R*^(*i*)^) := *A*_*i*_(*A*_*i−*1_, Ω).

The group sampling of *I*_1:*T*_ would be high dimensional and hard to transfer in the iteration of Metropolis-Hastings step, while the sequential sampling of *I*_1:*T*_ would be less efficient for the convergence of Markov chain [2, 4]. We utilized the mixture of MCMC kernels [2] to combine these two sampling strategies for the trade-off between the acceptance rate and the convergence of MCMC.

For the prediction of epidemic, we could use the information of the posterior distribution *π*(Ω, *I*_1:*T*_ |*H*_1:*T*_, *R*_1:*T*_, *N, β*). In Bayesian framework, since Ω is assumed to be random, *A*_*s*_(*T*) := *A*_*s*_(*A*_*T*_, Ω) is also random for given *A*_*T*_, *s > T*. Note that:

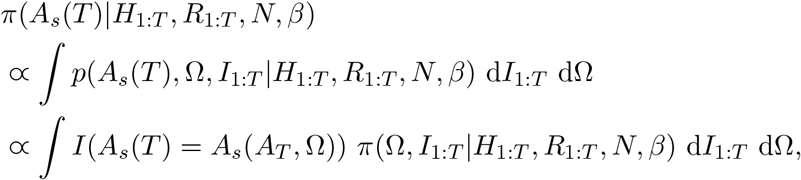

Hence the posterior distribution of *A*_*s*_(*T*) exists and could be calculated by *π*(Ω, *I*_1:*T*_ |*H*_1:*T*_, *R*_1:*T*_, *N, β*).

We applied *π*(*A*_*s*_(*T*)|*H*_1:*T*_, *R*_1:*T*_, *N, β*) for the predicted distributions of *A*_*s*_. The point estimation of parameters Ω, latent variables *I*_1:*T*_ and the predicted *A*_*s*_, *s > T* were presented as the median of the posterior distribution while 95% credible intervals were constructed with 2.5% and 97.5% quantiles.

#### 2.4. The mode migration

For simulating the epidemic trend of other countries, we applied the Bayesian SIHR model where *R*_*t*_ is time-invariant, i.e., *d* and *m* were chosen as 0 and + *∞*, respectively, for South Korea, Italy and the United States.

We used the real-time epidemic data in South Korea, Italy and the United States and estimated the posterior distributions of parameters, predicted numbers of cumulative confirmed cases and the missing numbers of infected and infectious individuals without isolation country by country. And then we migrated the estimated intensity of policy intervention of Wuhan, Wenzhou, and Shenzhen to South Korea, Italy, and the United States, i.e. changed the parameters *m* of South Korea, Italy, and the United States to the point estimation of *m* of Wuhan, Wenzhou, and Shenzhen, and simulated the epidemic trend, respectively. The point estimates were presented as the median of the posterior distribution while 95% credible intervals were constructed with 2.5% and 97.5% quantiles.

### 3. Real data analysis of COVID-19

#### 3.1. The estimation of policy patterns in China

To describe the effects of interventions in different cities in China, we used the observed data of Wuhan, Wenzhou and Shenzhen, to estimate the trends of *R*_*t*_, which were shown in Figure 4.

**Figure 4.**
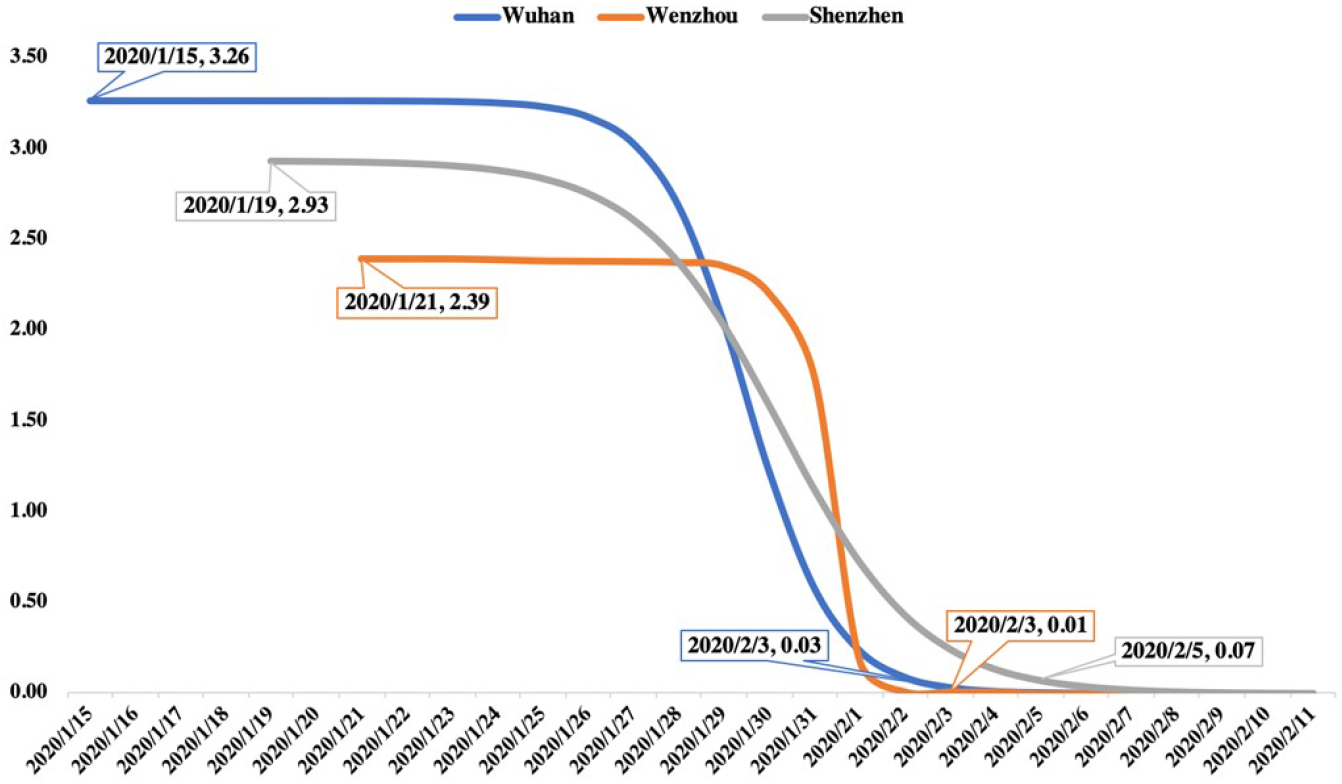
The trends of *R*_*t*_ for Wuhan, Wenzhou and Shenzhen.

The initial *R*_*t*_ for Wuhan was greater than that for Shenzhen, followed by Wenzhou. While Shenzhen was the city where the reproduction number started to decrease earliest, followed by Wuhan and Wenzhou. Moreover, the decreasing speed of *R*_*t*_ in Wenzhou was faster than that in Wuhan, followed by Shenzhen.

In SIHR model, the estimated *R*_*t*_ could reasonably reflect the patterns of policy interventions. From Figure 4, the different decreasing speeds of *R*_*t*_ implies different intensities of policy interventions. The intensity of policies for Wenzhou was the strongest, as opposite to be the weakest in Shenzhen of three cities. Comparing with details in Figure 1, Shenzhen issued relatively mild polices in the early period of outbreak, such as suspending entertainment places and tracking on the close contacts of confirmed cases, unlike the stringent policy interventions in Wenzhou, such as block both long-distance and short-distance social contacts. Indeed, mild strategy would cause less economic damage, oppositely, stringent strategy was used for the quick control of infectious disease.

#### 3.2. Simulation of the migration of policy patterns

The simulation of COVID-19 trend for constant *R*_*t*_ and time-varying *R*_*t*_ with mode migration of three policies of China in South Korea, Italy, and the United States are given as follows (Figures 5-7).

**Figure 5.**
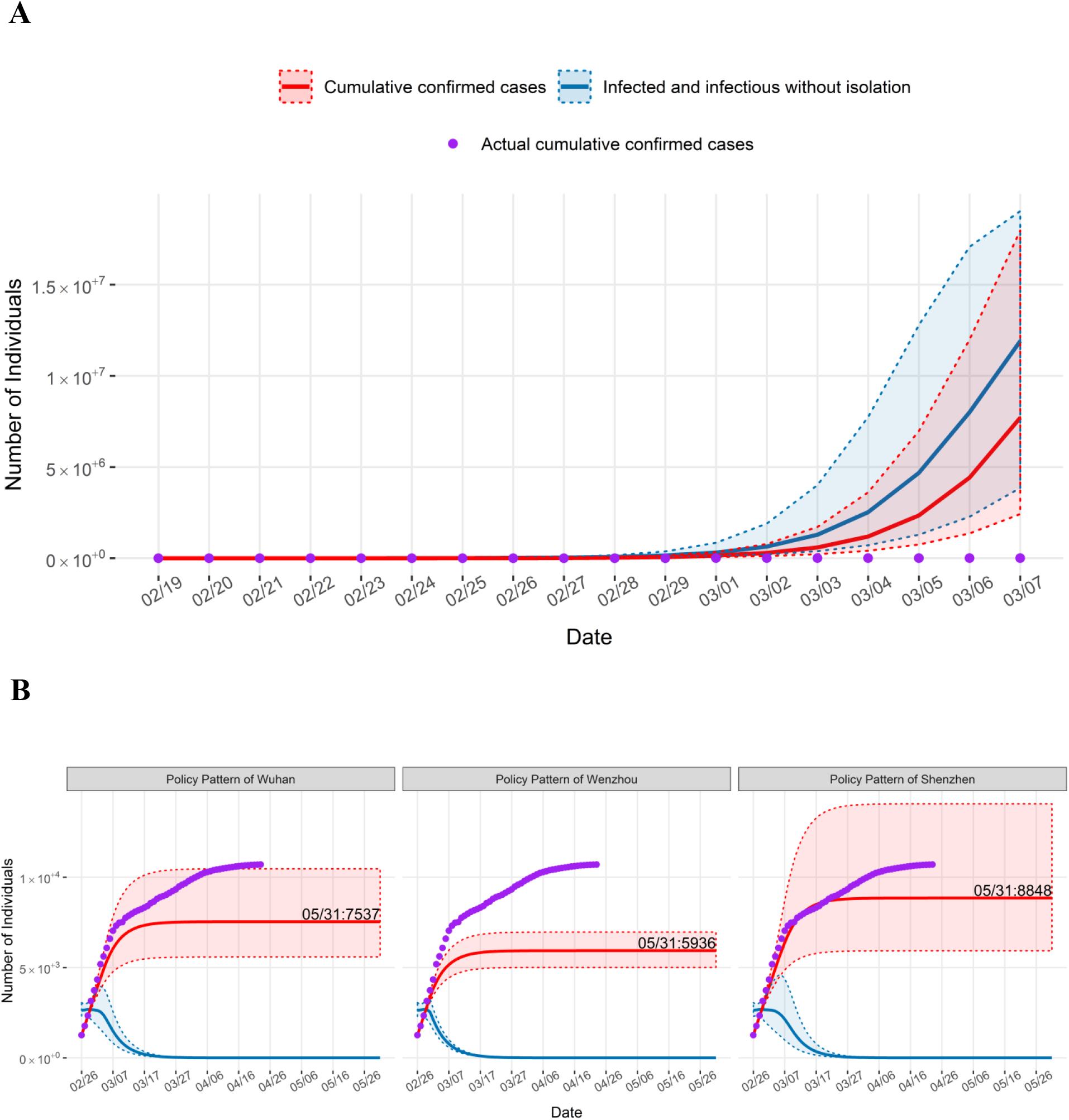
The simulation of COVID-19 trend for constant *R*_*t*_ (A) and time-varying *R*_*t*_ (B) of three policies in South Korea. The red curve with corresponding dashed lines represents the expected cumulative confirmed cases with 95% CI and the blue curve with corresponding dashed lines represent the expected number of infected and infectious without isolation (I) with 95% CI.

From our simulation, if interventions were not implemented since the starting date of our extracted data in South Korea, Italy and the United States, the expected cumulative confirmed cases would be 7705971 with corresponding 95% credible intervals (CI) (2419211,17969284) on March 7 (Figure 5A), 6680363 with 95% CI (442494,35236491) on March 29 (Figure 6A), and 34179685 with 95% CI (5274919,136469414) on April 17 (Figure 7A), respectively, when the expected cumulative confirmed cases would be over 10% of the total population in the corresponding country for the first time.

**Figure 6.**
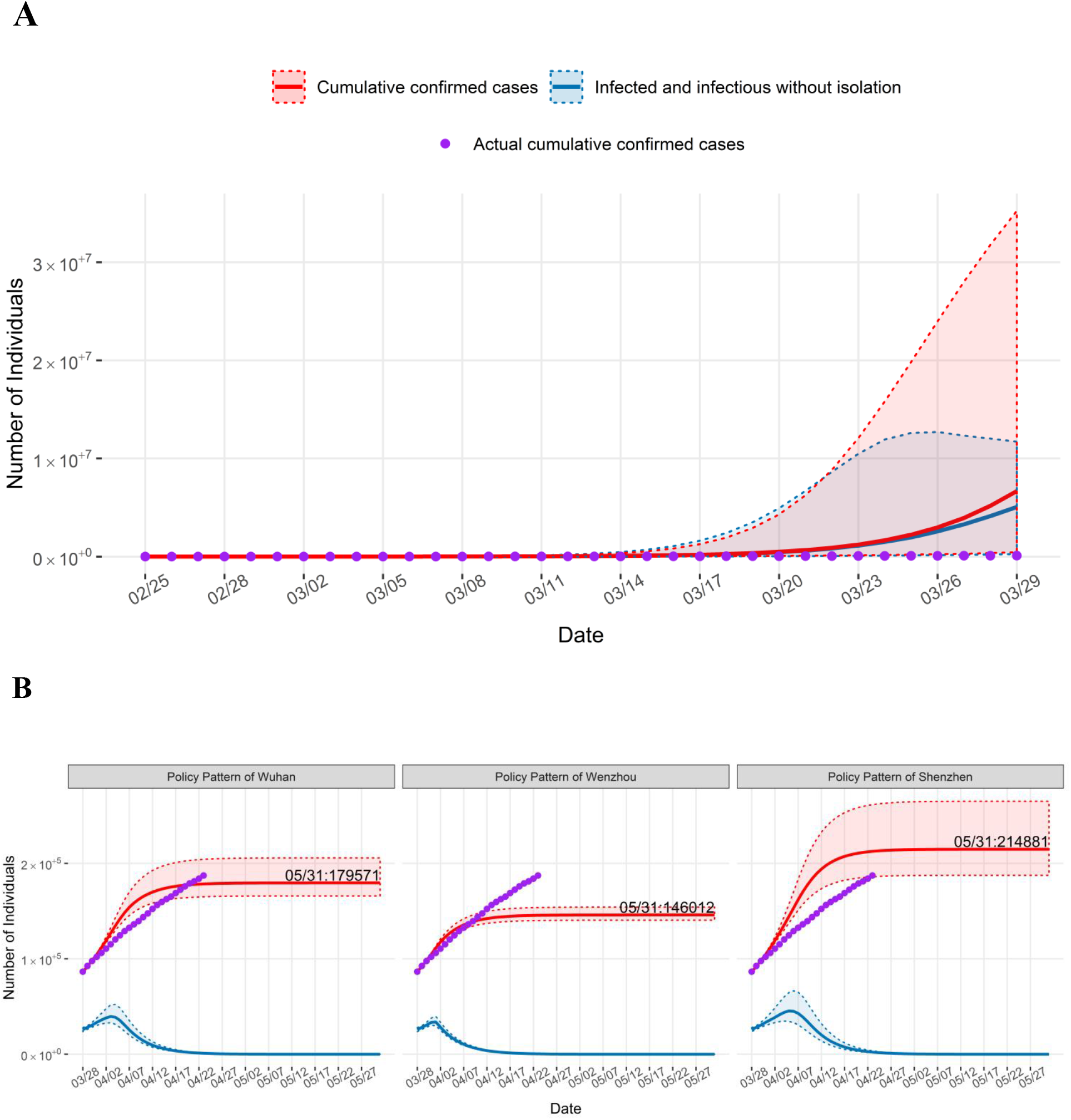
The simulation of COVID-19 trend for constant *R*_*t*_ (A) and time-varying *R*_*t*_ (B) of three policies in Italy. The red curve with corresponding dashed lines represents the expected cumulative confirmed cases with 95% CI and the blue curve with corresponding dashed lines represent the expected number of infected and infectious without isolation (I) with 95% CI.

**Figure 7.**
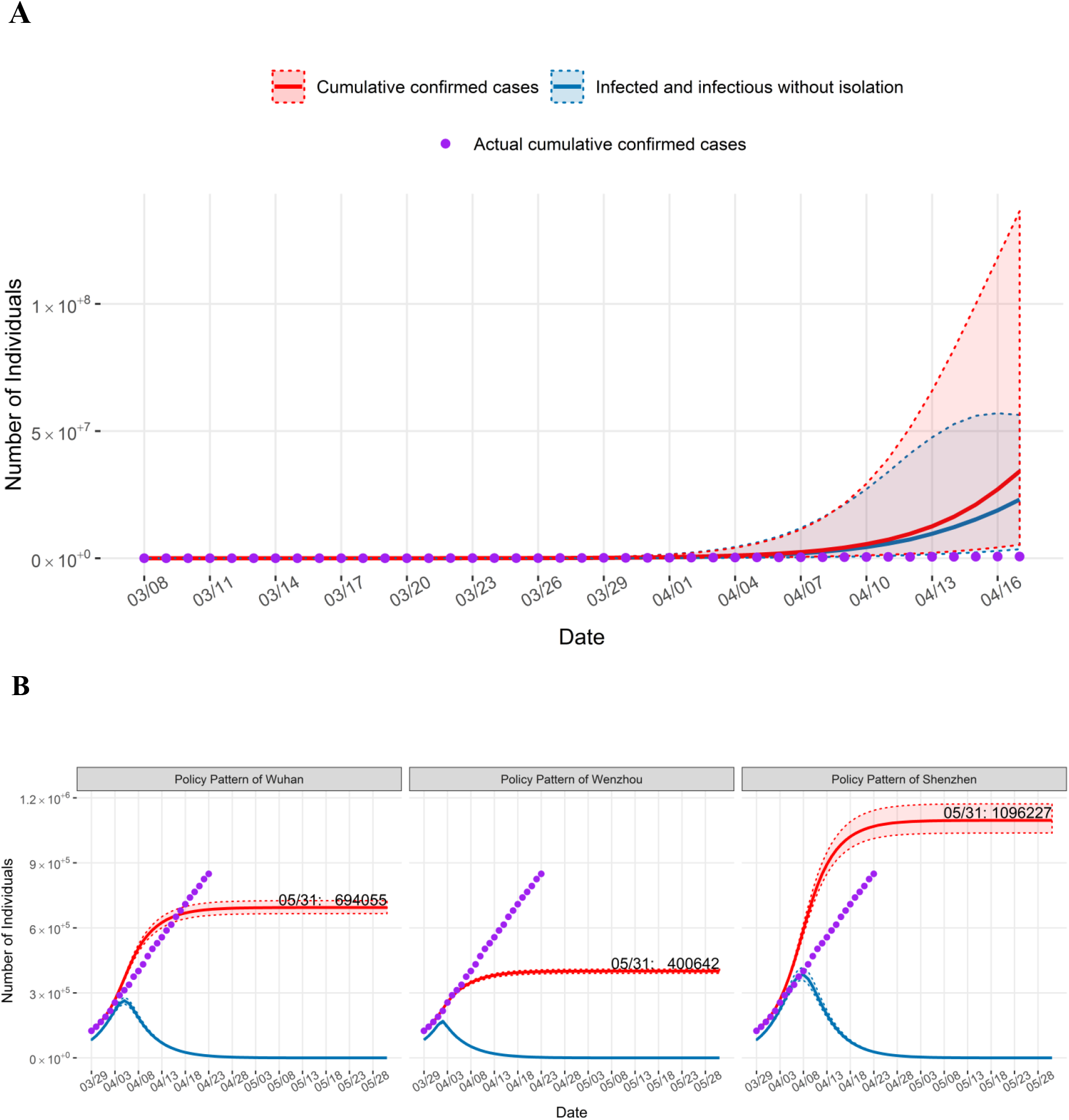
The simulation of COVID-19 trend for constant *R*_*t*_ (A) and time-varying *R*_*t*_ (B) of three policies in the United States. The red curve with corresponding dashed lines represents the expected cumulative confirmed cases with 95% CI and the blue curve with corresponding dashed lines represent the expected number of infected and infectious without isolation (I) with 95% CI.

**Figure 8.**
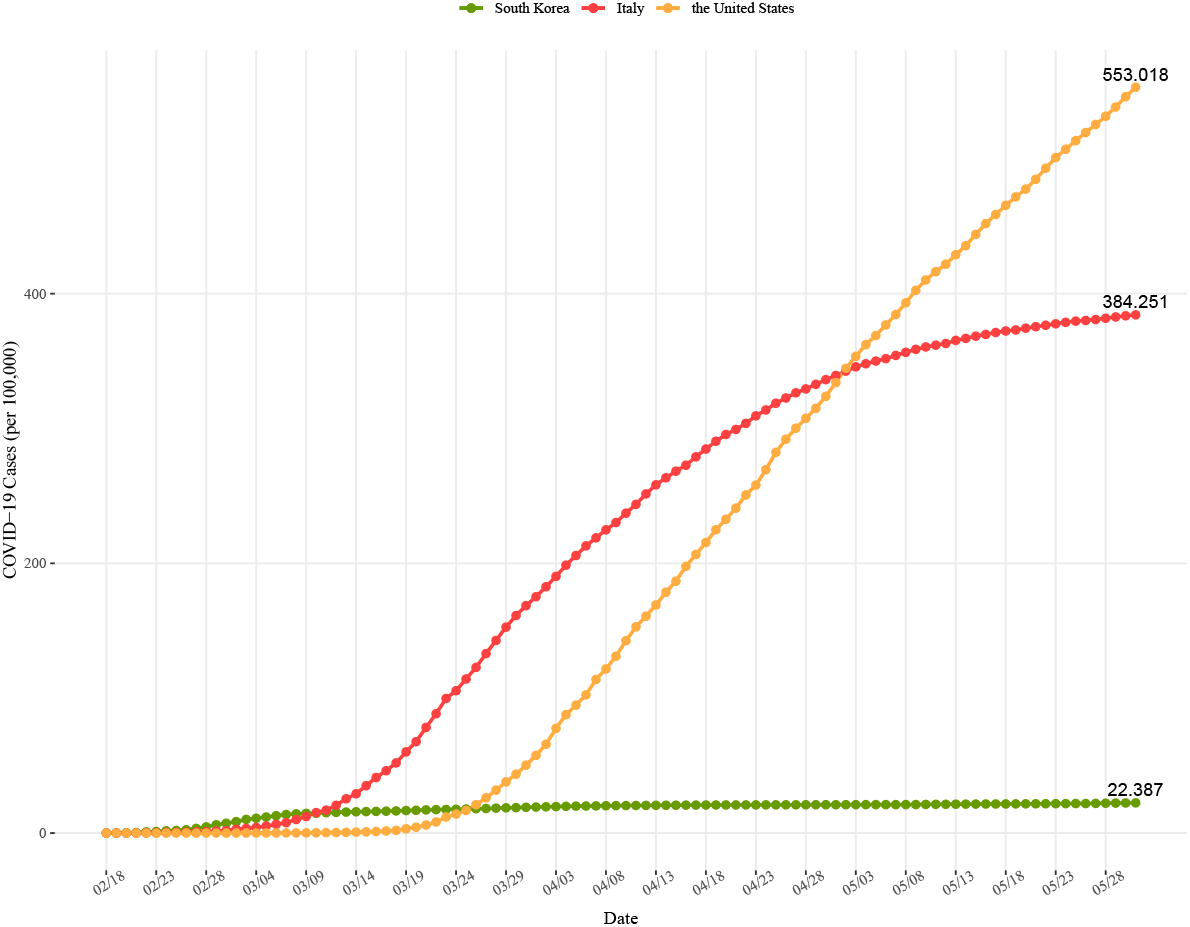
COVID-19 cases per 100, 000 in South Korea, Italy, and the United States from February 18 to May 31. On February 18, the 31st confirmed case in South Korea triggered the rapid spread in this country, so we used it as the starting date of wide spread outside mainland China.

As the estimated number of *I* was 2639 with corresponding 95% CI (2306,3055) on February 26 in South Korea, there is not much difference for the expected cumulative confirmed cases in the simulation of three urban interventions in South Korea (Figure 5B). However, for Italy, there is a substantial difference in the simulation of three policies implemented (Figure 6B). It may be caused by the large estimated number of *I* (25768) on March 28, which implies that it is cautious to carry out a policy at this stage of epidemic. Similarly, in the United States, there is a considerable difference from the policy of Shenzhen compared with other two patterns. One reason for this distinction is that the estimated number of *I* was 83232 (Figure 7B) on March 29, which implied certain risks for long-term outbreak of epidemic.

#### 3.3. Discussion

For South Korea, Italy and the United States, if interventions were not implemented, it can be expected that how tremendously large populations would be infected in these countries. Thus, the interventions are critical for the control of the fast spread of infectious disease.

Based upon our simulation, the actual cumulative confirmed cases of COVID-19 in South Korea from February 26 to March 9 were within the ranges of expected cumulative confirmed cases for Shenzhen pattern, whose intensity was relatively mild compared with both Wuhan and Wenzhou patterns. According to the report [19], South Korea has implemented similar interventions to Shenzhen pattern, where it implemented policies closely following and isolating the close contact individuals of confirmed cases to reduce the transmission of COVID-19. This indicated that the mitigation policies like Shenzhen pattern, implemented at an early stage of epidemic could effectively curb the outbreak of infectious diseases.

On the other hand, note that the expected number of cumulative confirmed cases under the interventions of Wenzhou for Italy would be the smallest. Comparing the actual situations of Italy, the cumulative confirmed cases were 187,327 on April 23, 2020, which were 28.7% higher than the simulation of COVID-19 under interventions of Wenzhou. If taking such highly stringent interventions, the epidemic situation could be mitigated where the expected number of cumulative confirmed cases would be 146,012 on May 31, decreasing by 19% and 32% compared to the implementation of policy of Wuhan and Shenzhen, respectively. It showed that containment policies like Wenzhou pattern were effective for quick control of the magnitude of outbreak of infectious disease.

In the United States, the cumulative confirmed cases were 849094 on April 23, 2020, and were 113.3% more than the simulation of COVID-19 under interventions of Wenzhou, which implied that the implementations of interventions of Wenzhou may significantly decrease the magnitude of the outbreak of COVID-19 for the United States. If taking the relatively mild interventions of Shenzhen, the situation would get worse and the expected cumulative confirmed cases would be over 1000000 in the United States on May 31.

## 4. Conclusion

In this paper, we proposed a novel compartmental epidemic model: Bayesian SIHR model, which took the pre-symptomatic permissibility of COIVD-19 into account and could estimate the policy patterns according to the real data, and further we can get reasonable assessments for the corresponding policy patterns through the mode migration between countries.

As a typical example, we used the epidemic data of three representative cities in China (Wuhan, Wenzhou and Shenzhen) and migrated the estimated policy modes to South Korea, Italy and the United States.

Our simulation results indicated that the mild interventions were difficult to control the magnitude of outbreak of infectious diseases when the number of infected and infectious without isolation (*I*) was quite large, but it was effective to implement at the early stage of epidemic. Once the epidemic became severe, the highly stringent interventions were needed to be implemented for the control of epidemic. Although it still takes time to make a reasonable assessment of the patterns and effects of these policies, the transmission patterns of COVID-19 under some typical intervention measures could be learned from them.

## Data Availability

The epidemic data were extracted for Wuhan, Wenzhou, and Shenzhen from the Chinese Center for Disease Control and Prevention (China CDC) (National Health Commission of the People's Republic of China, 2020), and the data for South Korea, Italy and the United States were downloaded through April 23, 2020 from COVID-19 Knowledge & Data Hub (Sciences, 2020).

http://2019ncov.chinacdc.cn/2019-nCoV/index.html

http://geodoi.ac.cn/covid-19/en/index.aspx

## Acknowledgements

We would like to thank all individuals who are collecting epidemiological data of the COVID-19 outbreak around the world.

## Supplemental material

### COVID-19 prevalence

Through May 31, 2020, the cumultative confirmed cases were 11,468 in South Korea, 232,664 in Italy and 1,819,788 in the United States, while their corresponding COVID-19 cases per 100, 000 were 22.387, 384.251 and 553.018 in South Korea, Italy, and the United States, respectively.

**Table 1.** COVID-19 cases per 100,000 in South Korea, Italy and the United States from February 18 to May 31.

| Date | South Korea | Japan | Singapore |
| --- | --- | --- | --- |
| 2/18/2020 | 0.061 | 0.005 | 0.005 |
| 2/28/2020 | 4.562 | 1.078 | 0.018 |
| 3/9/2020 | 14.598 | 12.261 | 0.174 |
| 3/19/2020 | 16.720 | 60.206 | 3.198 |
| 3/29/2020 | 18.708 | 152.720 | 37.946 |
| 4/8/2020 | 20.271 | 224.875 | 121.723 |
| 4/18/2020 | 20.796 | 284.779 | 215.470 |
| 4/28/2020 | 20.990 | 329.337 | 307.583 |
| 5/8/2020 | 21.126 | 356.495 | 393.207 |
| 5/18/2020 | 21.601 | 372.312 | 465.482 |
| 5/28/2020 | 22.145 | 381.732 | 531.555 |
| 5/31/2020 | 22.387 | 384.251 | 553.018 |

Comparing three countries, we can find that South Korea had the highest COVID-19 cases per 100,000, followed by Italy, and the United States had the lowest from February 18 to March 9, 2020. On March 10 the COVID-19 cases per 100,000 in Italy surpassed that in South Korea, rising rapidly. After May, the United States had the largest COVID-19 cases per 100,000.

